# Vitamin D and changes in cognitive performance in mid and later life: findings from the English Longitudinal Study of Ageing (ELSA)

**DOI:** 10.1101/2022.11.14.22282290

**Authors:** Sergiu Grajdean, Cesar de Oliveira, Dorina Cadar

## Abstract

**Background:** Vitamin D has been found to be protective against biological processes associated with Alzheimer’s disease and cognition, including amyloid-β deposition, but the evidence in relation to cognitive decline is scarce.

**Objective:** This study aims to investigate the relationship between 25(OH)D serum levels and changes in cognitive performance over time in middle-aged and older English adults.

**Research Design and Methods:** An observational study design. We analyzed data collected biannually over a 6-year follow-up from a prospective cohort, the English Longitudinal Study of Ageing (ELSA). Mixed Linear Models analyses were conducted. The analytical sample consisted of 5,365 participants aged 50 and older without a diagnosis of dementia at baseline. Cognitive performance was ascertained with memory and orientation. Both tests were administered at every ELSA wave from Wave 6 (2012-2013) to Wave 9 (2018-2019). The values of blood levels of 25(OH)D and all covariates were considered at Wave 6 since this was the first time 25(OH)D was assessed. Blood levels of 25(OH)D were dichotomized in high versus low using the common threshold of 31nmol/l.

**Results:** A significant positive association was found between low serum levels of 25(OH)D and baseline memory scores (β= -0.28, SE=0.05, p≤0.001), but not with changes in memory over time. Furthermore, baseline blood levels of 25(OH)D were not associated with baseline orientation or changes in orientation over time.

**Discussion:** In this representative sample of the English population, we found that lower 25(OH)D serum levels were associated with a lower baseline memory but did not influence the change in cognitive functioning over time.

**Implications:** This study provides further clarification in understanding the deficient role of 25(OH)D on cognitive performance and the change in cognitive functioning over time.

## Introduction

Vitamin D is essential for the optimal function of the cardiovascular, endocrine, and nervous systems (1). The primary sources of Vitamin D are diet, supplements, and ultraviolet light synthesized in the skin (2). Vitamin D insufficiency, hypovitaminosis D, is widespread worldwide, becoming a significant public health issue (3). The best biomarker of vitamin D in our organisms is currently thought to be the serum level of 25-hydroxyvitamin D, 25(OH)D. Serum levels of 25(OH)D decrease with age, leading to an increased risk of developing different diseases (4). Older adults present lower 25(OH)D blood levels and are at higher risk of developing diseases associated with hypovitaminosis D, such as osteoporosis, cardiovascular diseases, hypertension, cancer, and type II diabetes (5).

While specific roles of blood levels of 25(OH)D have been well established and supported by research, the role of blood levels of 25(OH)D in cognitive performance still needs to be clarified, with different results from cross-sectional and longitudinal studies (6). It has been found that areas involved in cognitive functions such as memory, executive functions, information processing, and concentration have receptors via which 25(OH)D exerts its actions (7). A review of published and unpublished controlled trials on the effect of Vitamin D on cognition, including memory, executive functions, information processing, and overall cognitive performance with participants aged 40 years and more with no cognitive impairment at the time of the experiments, did not find any significant association between higher levels of Vitamin D and cognition, both overall performance and separate domains of it such as memory or executive functions (8).

However, a systematic review of cross-sectional studies performed in older adults detected a significant positive relationship between low blood levels of 25(OH)D and low scores on tests that assessed global cognitive performance while failed to find any prospective cohort study to support this association in the long term (9). This is supported by another systematic review of cross-sectional studies that showed an inverse relationship. In contrast, individuals who obtained higher scores on tests on global cognitive performance presented higher blood concentrations of 25(OH)D (10).

Furthermore, emerging evidence indicates that an insufficient blood level of 25(OH)D could be one of the markers of cognitive decline, with cross-sectional studies suggesting that lower levels of 25(OH)D are associated with a steeper decline in cognitive performance (11). However, with limited longitudinal studies on the association between serum levels of 25(OH)D and cognitive decline, further investigation is required.

While the relationship between blood levels of 25(OH)D and cognitive decline is still not clear, emerging evidence suggests an association between low blood levels of 25(OH)D and a higher risk of developing cognitive impairment and Alzheimer’s Disease. A recent meta-analysis indicated that blood levels of 25(OH)D less than 25nmol/l might contribute to an increased risk of developing dementia (12). Cross-sectional studies have found that participants with cognitive impairment and Alzheimer’s Disease have lower levels of 25(OH)D than healthy individuals (7). However, compared with cross-sectional studies, the longitudinal evidence on the association between serum levels of 25(OH)D and the incidence of cognitive impairment and Alzheimer’s disease have reported mixed results and a less powerful association (11).

The previous results from both cross-sectional and longitudinal studies on the relationship between blood levels of 25(OH)D and cognition are mixed. However, emerging evidence suggests that 25(OH)D plays a role in cognitive health. Some animal studies have found that induced insufficient blood levels of 25(OH)D in young rats led to structural modifications in the brain cortex, lateral ventricles, and their behavior (13). Moreover, neuroimaging studies have suggested similar results as those found in the studies conducted on animals. For example, it has been found that individuals with insufficient blood levels of 25(OH)D presented larger lateral ventricles than individuals with normal blood levels of 25(OH)D (6). It has also been suggested that 25(OH)D also plays a role in neurogenesis (birth of new neurons), in the expression of various neurotrophic factors, and in maintaining the calcium equilibrium, slowing down neurodegeneration (14). Other studies indicate that 25(OH)D is involved in the regulation of nerve growth factor release, which is crucial for the survival of neurons in the hippocampus, an important area for memory (14). Research has also suggested that 25(OH)D might be involved in detoxification and β-amyloid removal, playing a preventive role in the onset of Alzheimer’s Disease (15). This was supported by a study conducted in 2016 on humans that indicated that 25(OH)D contributes to decreasing brain β-amyloid (16).

The inconsistencies observed in the literature so far might be attributed to several factors, such as extreme heterogeneity of the studies’ methodologies, different definitions of optimal levels of blood levels of 25(OH)D, various tests of cognitive performances ability employed measuring other areas of cognition and numerous confounders that cannot always be controlled by researchers. Moreover, there is a lack of longitudinal studies investigating the relationship between blood levels of 25(OH)D and cognitive performance in adults aged 50 or above free of cognitive impairment or dementia. Therefore, there is a considerable gap in the current literature regarding the longitudinal studies investigating the relationship between serum levels of 25(OH)D and cognitive performance in older adults without a diagnosis of dementia.

Therefore, the present study aims to fill the gap in the existing literature by examining the associations between blood levels of 25(OH)D and changes in cognitive performance (memory and orientation) over a 6-year follow-up using a representative sample of older English adults. We hypothesise that lower 25(OH)D serum levels would be associated with lower baseline cognitive performance and faster decline over time in each of the two cognitive domains (memory and orientation).

## Method

### Study population

This is an observational study design. Data came from the longest-running national representative sample of the English population aged 50 or older, the English Longitudinal Study of Aging (ELSA) (17). The study started in 2002, with nine waves of data collection conducted biennially. A more detailed description of this multidisciplinary observational panel can be found elsewhere (17).

For the present investigation, the baseline information regarding the serum levels of 25(OH)D, cognitive performance, and covariates were taken from Wave 6 (2012-2013) since it was the first time when 25(OH)D concentrations were collected in ELSA. Participants who had a history of convulsions, bleeding disorders such as haemophilia, or were under current anticoagulant treatment and those who did not give consent were excluded from blood sampling.

The six years of follow-up information regarding the change in cognitive performance was considered from Wave 6 (2012-2013) to the latest wave 9 (2018-2019). Participants who presented dementia at wave 6 were excluded from the current analysis. For these analyses, only core members aged 50 years and older at baseline (wave 6) were included. For more information, see the flow chart of the analytical sample selection in Figure 1.

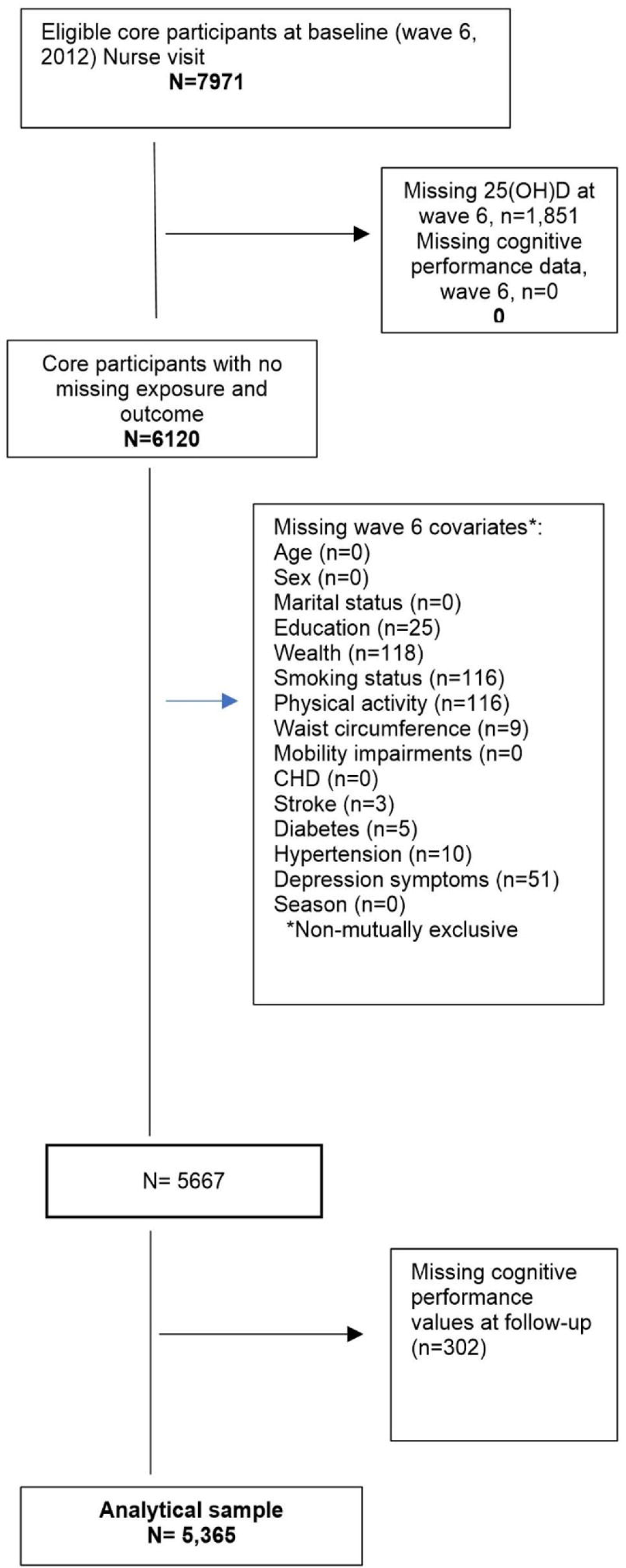

### Exposure: Blood levels of 25(OH)D

Blood levels of 25(OH)D were collected as part of the health examination, i.e., nurse visit at wave 6, the first wave that included assessment of blood levels of 25(OH)D, using the DiaSorin Liaison® immunoassay. This method allows us to analyse the total circulating of 25(OH)D, detecting both 25(OH)D_2_ and 25(OH)D_3_. Serum 25(OH)D levels were classified into two groups: low blood levels of 25(OH)D < 31nmol/l and high blood levels of 25(OH)D ≥ 31nmol/l, the latter used as a reference group in the current analysis. The threshold of 31nmol/l was chosen based on the existing literature that states that most studies consider levels under 31nmol/l to be deficient (18).

### Outcome: Cognitive performance and change in cognitive performance over time

We assessed the amnestic and non-amnestic domains of cognitive performance. The amnestic domain was assessed with verbal memory by a word list learning test, which consists of immediate and delayed recall of a random list of 10 words. After listening to 10 words, the participants were asked to repeat as many words as they could remember immediately and after a short delay during which they performed other cognitive tasks. The total possible memory score was 20, with higher scores representing a better memory performance.

The non-amnestic domain was assessed by an orientation test. The scores of this test varied from 0 to 4, with the higher the score, the better the orientation performance. For the changes in cognitive performance over time analysis, the memory and orientation scores were considered from the baseline (wave 6) to wave 7, 8, and 9, i.e., 6 years follow-up. One of the main reasons for choosing these two cognitive domains was that the scores were not based on self-assessment but on valid, objective tests.

### Covariates

The following socio-demographic and physical health covariates were included: age, sex, marital status, education, wealth, smoking status, physical activity, waist circumference, mobility impairments, coronary heart disease, stroke history, diabetes, hypertension, symptomology of clinical depression, and the season when the serum 25(OH)D was collected.

Age was centred, subtracting the mean from the actual baseline age variable, and the models were adjusted for sex. Marital status was included as a binary variable describing whether participants were unmarried or married at baseline. Education was classified into three groups: no qualification, intermediate (including those who completed O levels or the equivalent and those who completed A levels or the equivalent such as vocational qualifications), and higher (including those having a university degree or higher). Socioeconomic position was assessed using the total non-pension household wealth, including savings, investments, and property wealth, divided into quintiles. Smoke was included in the analysis as a binary variable, i.e., non-smokers and smokers. The levels of physical activity were obtained from the self-report questionnaire, which aimed at establishing the frequency of engaging in physical exercise, and participants were divided into two categories: those who did none or rarely and those who reported having been engaging in moderate to vigorous physical activity. It has been suggested that waist circumference could influence cognitive performance (19). In the current analysis, waist circumference was divided into three groups: low, medium, and high. As mobility impairment could influence 25(OH)D levels as it might impact direct exposure to sunlight UVBs, participants were divided into those with no mobility impairment and those with one or more mobility impairments. Coronary heart disease (CHD) history was also considered. It was transformed into a binary variable with participants divided into those who have never had it and those with a history of CHD. Stroke, diabetes, and hypertension were also used as binary variables, distinguishing between those who had never had these conditions and those with a prior history at baseline. Depressive symptoms were measured using the short version of the CES-D scale (20), and the sample was dichotomized into participants who had never experienced them (CES-D < 4) and those who had a history of depressive symptoms at wave 6 (CES-D >= 4). The analysis was adjusted for the time of year (seasons) when the serum 25(OH)D was collected to adjust for the variation in the synthesis of 25(OH)D according to sunlight UVB exposure: winter, spring, summer, and autumn.

### Statistical Analysis

Linear mixed effects models (LMMs) were conducted to investigate whether there was an association between low or high blood levels of 25(OH)D and changes over time of memory and orientation performance during the follow-up period, i.e., waves 6 to 9. LMM consider the fact that repeated measures of both memory and orientation in the same participants are correlated. Both random intercepts and random slopes were used. A “time” variable was created in the current analysis to indicate the follow-up from wave 6 to 9, with every unit representing a 2-year increase in the follow-up time. Memory and orientation change was modeled as a linear function of time measured from the baseline wave until the end of the study period. Random effects for the intercept and slope were fitted for each individual, allowing participants to have different scores at baseline and rates of change in memory. To test whether cognitive trajectories differed between participants with low and high vitamin D, we included in the model vitamin D, covariates, time, time×vitamin D, and time×covariates. Unstandardized coefficients and standard errors (SE) for baseline cognition (intercept) and linear change (slope) were presented for each cognitive test (memory and orientation) from fully adjusted models. Missing observations were assumed to be missing at random (MAR), and model assumptions were verified by examining residuals computed from the predicted values. LMMs offer a simple alternative to handle missing data under MAR without requiring imputations, so we used fully observed data. Baseline cross-sectional sample weights were used to ensure that the sample is representative of the general population. Analyses were performed for memory and orientation, respectively, using STATA 16.1 version (StataCorp LP). The standard level of statistical significance of 0.05 was considered. The manuscript was written following the Strengthening the Reporting of Observational Studies in Epidemiology (STROBE) guidelines.

## Results

### Study population characteristics

Table 1 summarises the characteristics of 5,365 ELSA participants at baseline, wave 6. Regarding their serum levels of 25(OH)D, 84.36% of the sample presented blood levels equal to or higher than 31 nmol/l, while 15.64% had levels of 25(OH)D less than 31 nmol/l. The lowest concentration in this analytical sample was 9 nmol/l. As mentioned before, the total possible score of memory was 20, with higher scores representing a better memory performance. In the current study, the mean memory score was 11.01 with a standard deviation of ±3.39, whilst the orientation score had a mean of 3.81 (out of the highest score of 4) with a standard deviation of ±0.47. The mean age of the included participants was 67.12 with a standard deviation of ± 8.52 years, and 55.12% were women. More than half of them (66.64%) were married at the baseline. Most participants (78.21%) had an intermediate or higher education level, and more than half of them (66.70%) fell within the top three quintiles for total wealth. The great majority of the participants (88.70%) were non-smokers, and 81.81% of them claimed to do moderate or vigorous physical exercise. Slightly more than half of the subjects (51.44%) had a high waist circumference. Regarding mobility impairments, more than half, 51.11% of the included participants reported having one or more. However, concerning other physical conditions that our participants declared to have, a large proportion (92.45%) did not have any history of coronary heart disease, 96.18% did not have a stroke, and 89.60% of them were not diabetics. However, almost half of the subjects, 44.27%, had reported suffering from hypertension. In terms of depressive symptomology, 88.50% of the participants reported not having any. Looking at the season when the blood samples were collected, 42.96% of the participants had their samples collected in the autumn. For a more detailed presentation of the study participants’ characteristics, see Table 1.

**Table 1.**
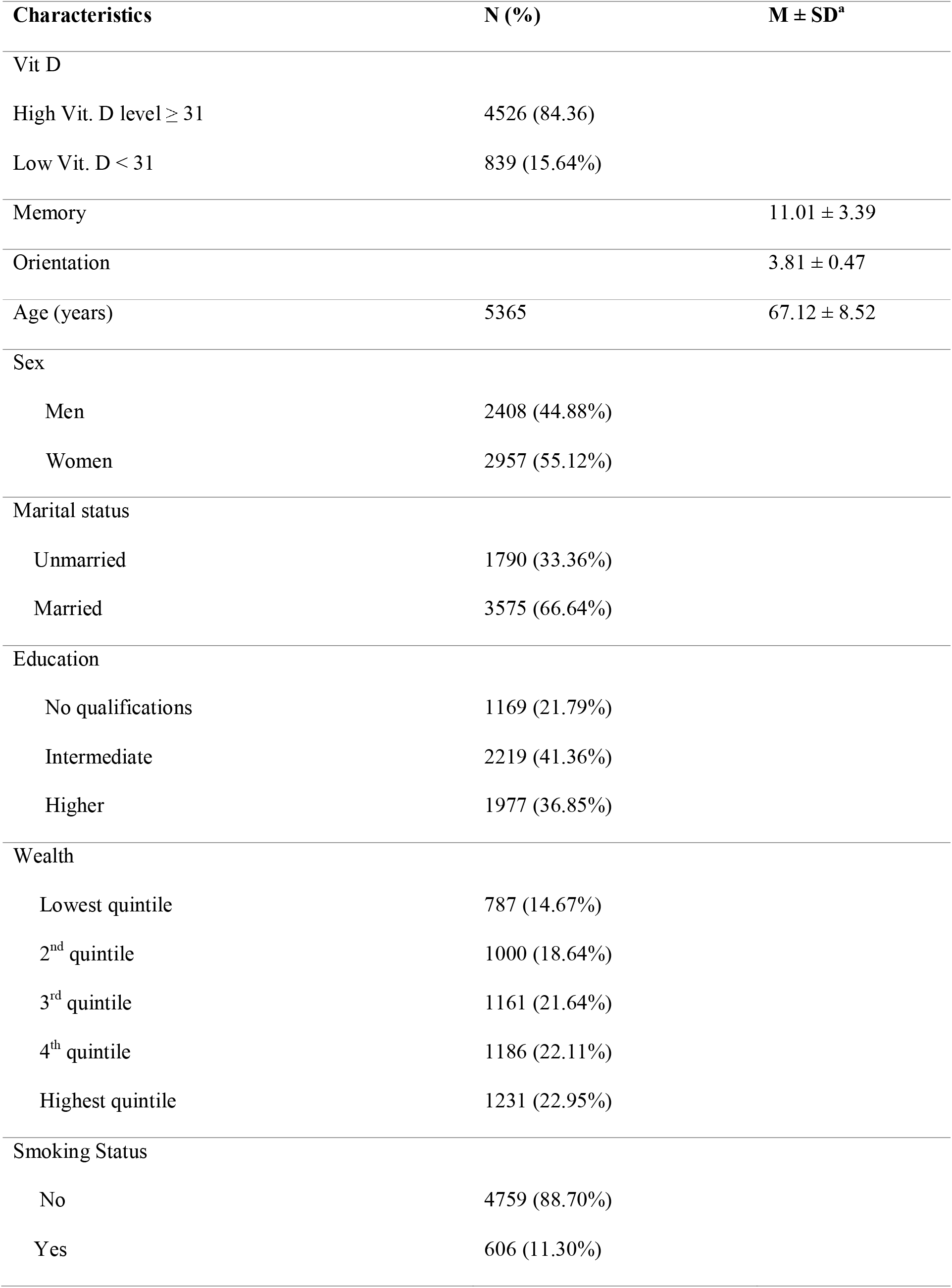
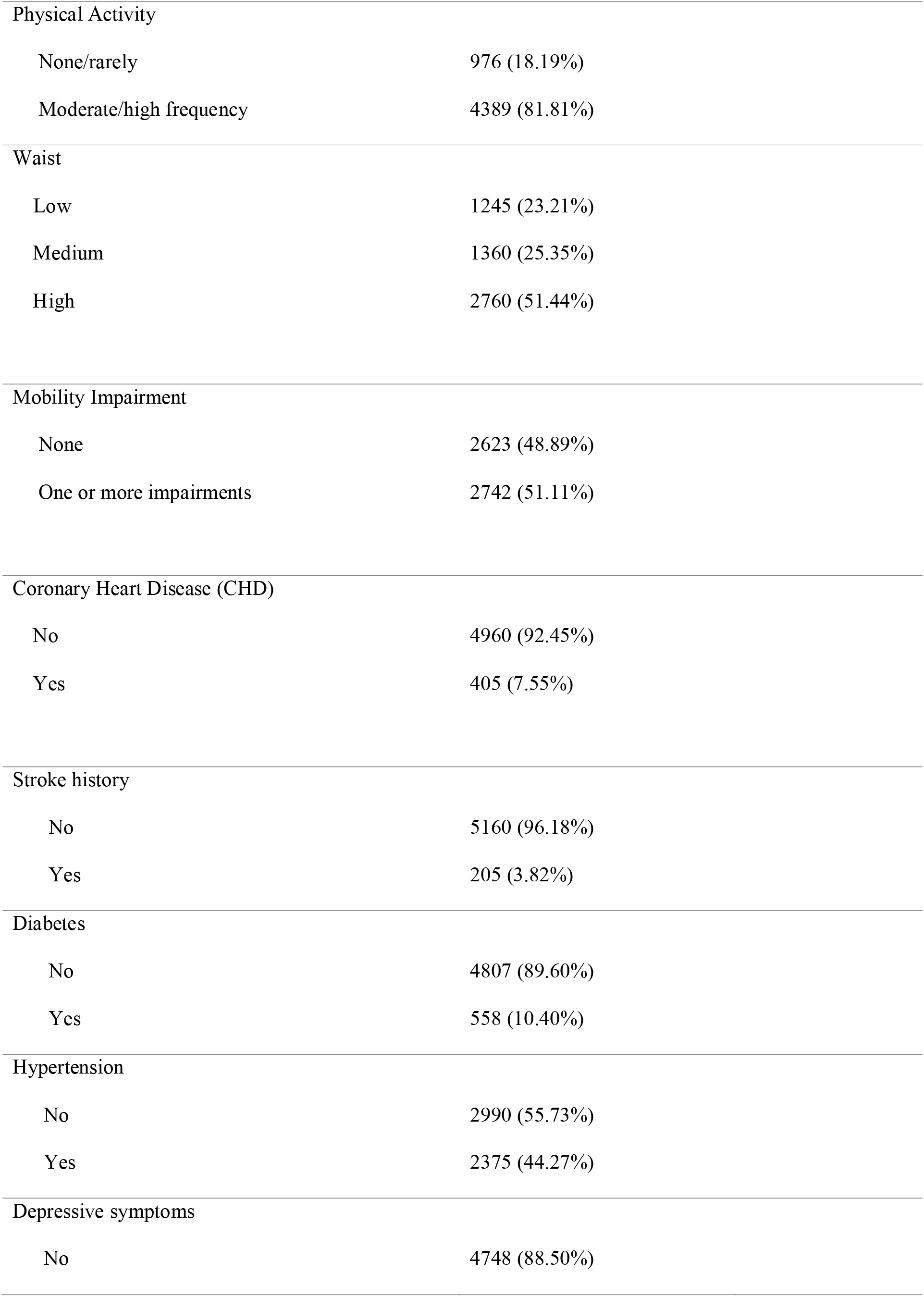
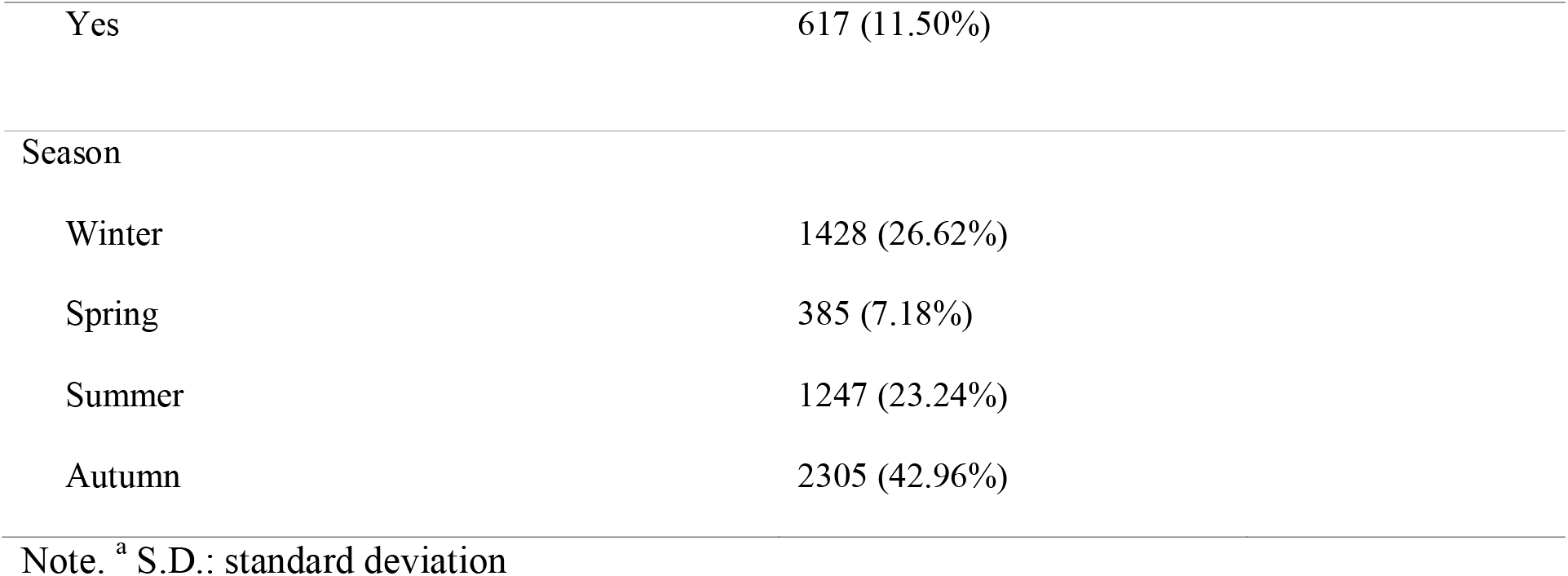
Characteristics of the study participants at baseline, Wave 6.

| Characteristics | N (%) | M ± SD<sup>a</sup> |
| --- | --- | --- |
| Vit D |  |  |
| High Vit. D level ≥ 31 | 4526 (84.36) |  |
| Low Vit. D < 31 | 839 (15.64%) |  |
| Memory |  | 11.01 ± 3.39 |
| Orientation |  | 3.81 ± 0.47 |
| Age (years) | 5365 | 67.12 ± 8.52 |
| Sex |  |  |
| Men | 2408 (44.88%) |  |
| Women | 2957 (55.12%) |  |
| Marital status |  |  |
| Unmarried | 1790 (33.36%) |  |
| Married | 3575 (66.64%) |  |
| Education |  |  |
| No qualifications | 1169 (21.79%) |  |
| Intermediate | 2219 (41.36%) |  |
| Higher | 1977 (36.85%) |  |
| Wealth |  |  |
| Lowest quintile | 787 (14.67%) |  |
| 2<sup>nd</sup> quintile | 1000 (18.64%) |  |
| 3<sup>rd</sup> quintile | 1161 (21.64%) |  |
| 4<sup>th</sup> quintile | 1186 (22.11%) |  |
| Highest quintile | 1231 (22.95%) |  |
| Smoking Status |  |  |
| No | 4759 (88.70%) |  |
| Yes | 606 (11.30%) |  |

|  |  |
| --- | --- |
| Physical Activity |  |
| None/rarely | 976 (18.19%) |
| Moderate/high frequency | 4389 (81.81%) |
| Waist |  |
| Low | 1245 (23.21%) |
| Medium | 1360 (25.35%) |
| High | 2760 (51.44%) |
| Mobility Impairment |  |
| None | 2623 (48.89%) |
| One or more impairments | 2742 (51.11%) |
| Coronary Heart Disease (CHD) |  |
| No | 4960 (92.45%) |
| Yes | 405 (7.55%) |
| Stroke history |  |
| No | 5160 (96.18%) |
| Yes | 205 (3.82%) |
| Diabetes |  |
| No | 4807 (89.60%) |
| Yes | 558 (10.40%) |
| Hypertension |  |
| No | 2990 (55.73%) |
| Yes | 2375 (44.27%) |
| Depressive symptoms |  |
| No | 4748 (88.50%) |

|  |  |
| --- | --- |
| Yes | 617 (11.50%) |
| Season |  |
| Winter | 1428 (26.62%) |
| Spring | 385 (7.18%) |
| Summer | 1247 (23.24%) |
| Autumn | 2305 (42.96%) |

Note. <sup>a</sup> S.D.: standard deviation

### Blood levels of 25(OH)D and change in memory over time

The associations between 25(OH)D blood levels and changes in memory performance over time are presented in Table 2. The results of the fully adjusted model indicate that ELSA participants had a mean intercept of 7.45, SE=0.25, p≤0.001 in memory, and the rate of linear change declined over time with an average of β= -0.28, SE=0.05, p≤0.001. Baseline low blood levels of 25(OH)D were positively associated with lower memory at baseline. This indicates that participants with lower blood levels of 25(OH)D < 31nmol/l presented a lower baseline memory score. Age (β= -0.12, SE= 0.01, p≤0.001), mobility impairment (β= -0.04, SE=0.02, p≤0.005), elevated depressive symptoms (β= -0.05, SE=0.13, p≤0.001), and season (β= -0.24, SE=0.11, p≤0.005), when the blood sample was collected were all related to poorer memory at baseline. On the other hand, being female (β= 1.19, SE=0.08, p≤0.001), having a higher level of education (β=1.68, SE=0.12, p≤0.001), being wealthier (β= 1.17, SE=0.15, p≤0.001 and doing physical exercise at a higher rate (β= 0.46, SE=0.12, p≤0.001) has been shown to be associated with higher memory scores. There was no statistically significant association between blood levels of 25(OH)D and changes in memory performance over time. Among the covariates, only education has been shown to be positively associated with slower memory performance decline over time (β= 0.07, SE=0.02, p≤0.01), and only age has been linked with a faster memory decline (β= -0.01, SE=0.00, p≤0.001).

**Table 2.**
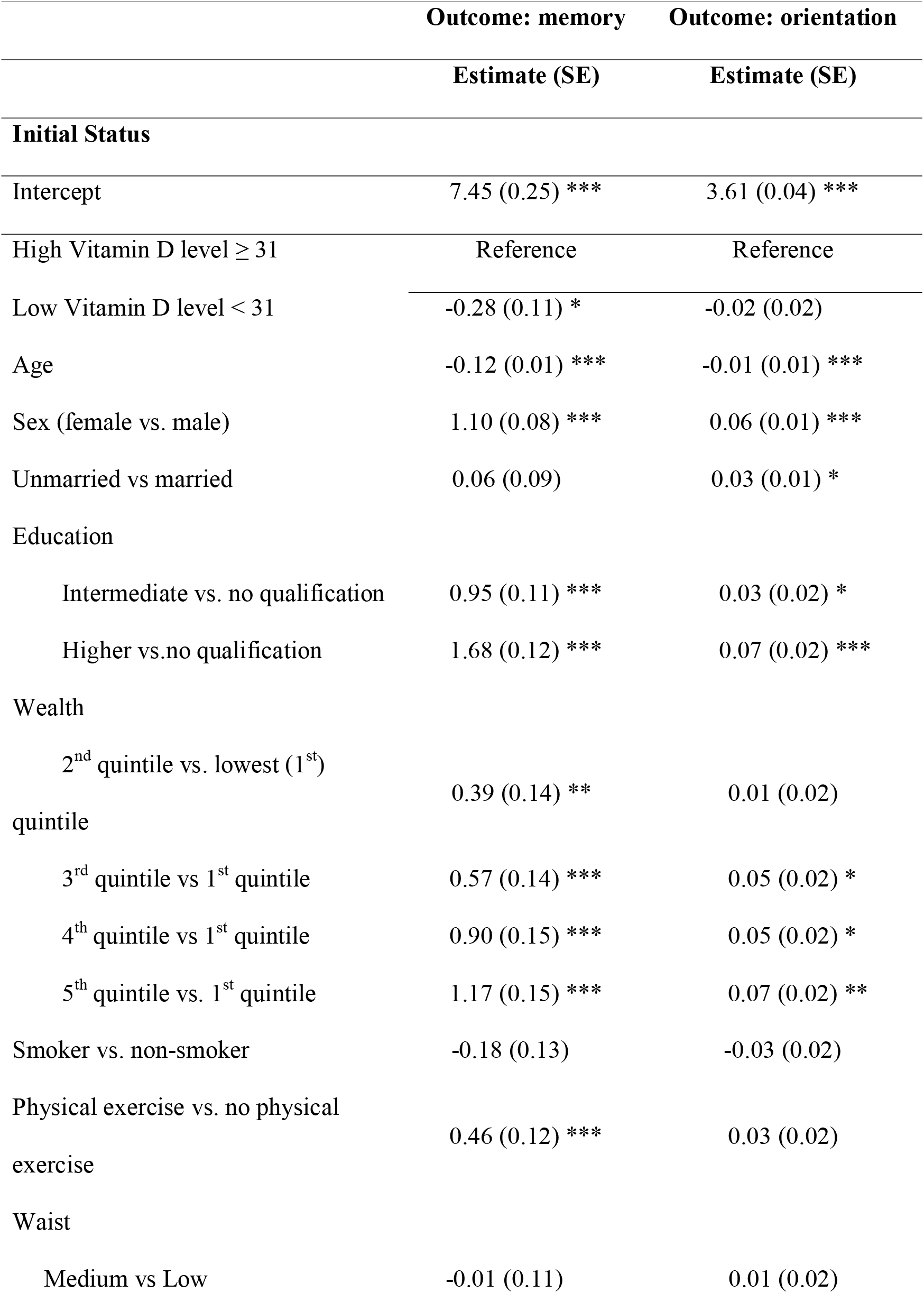
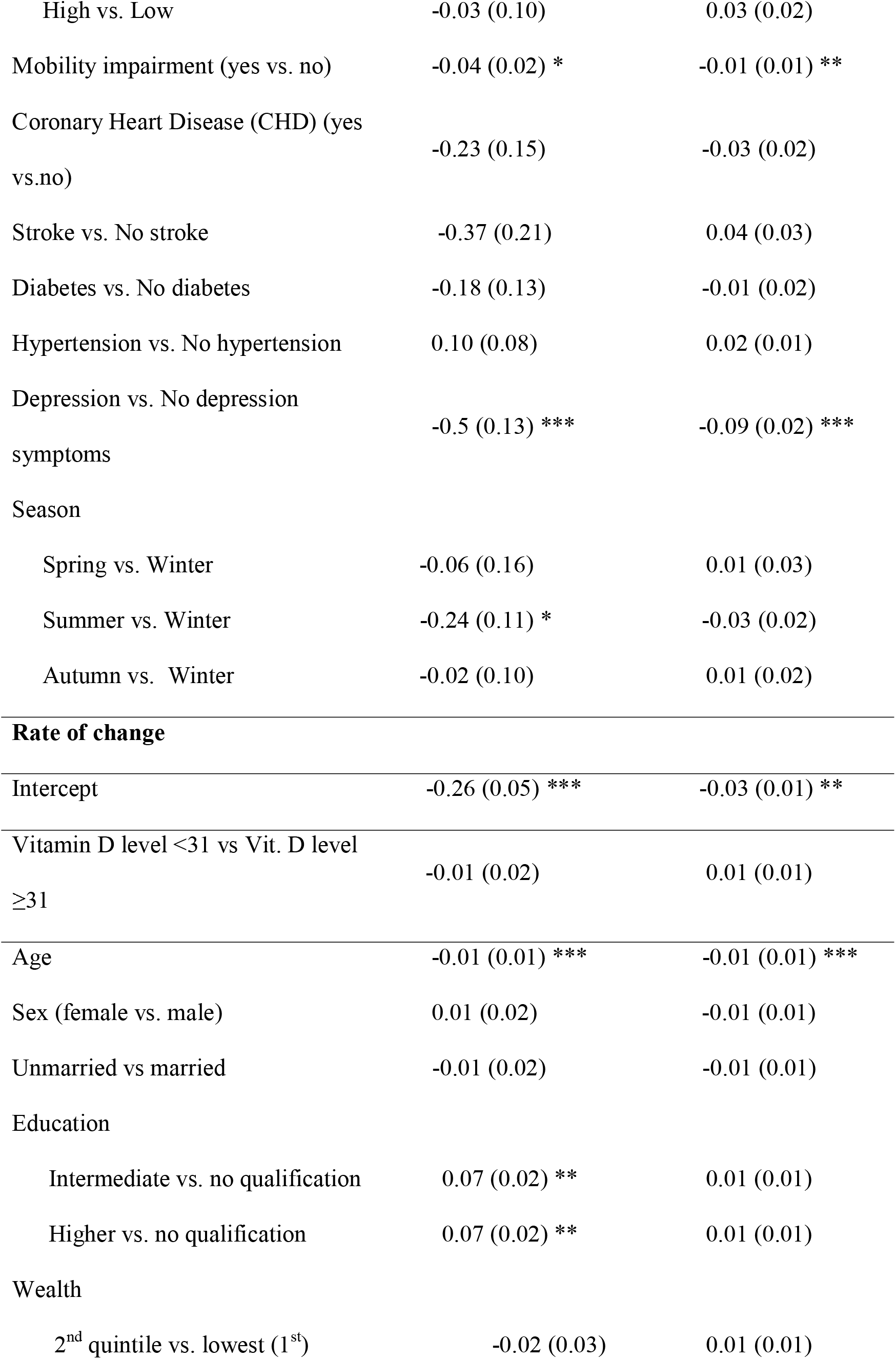
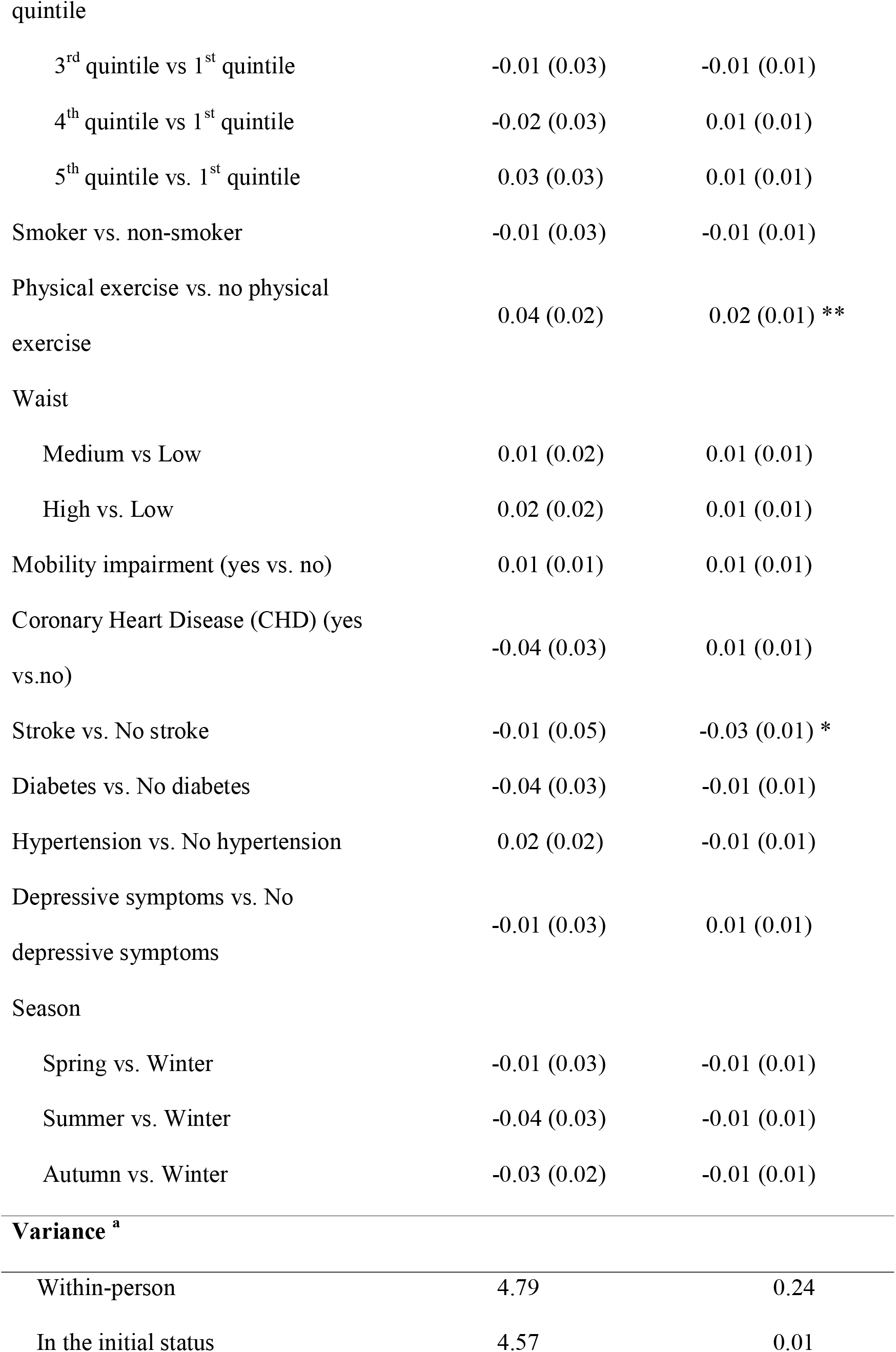
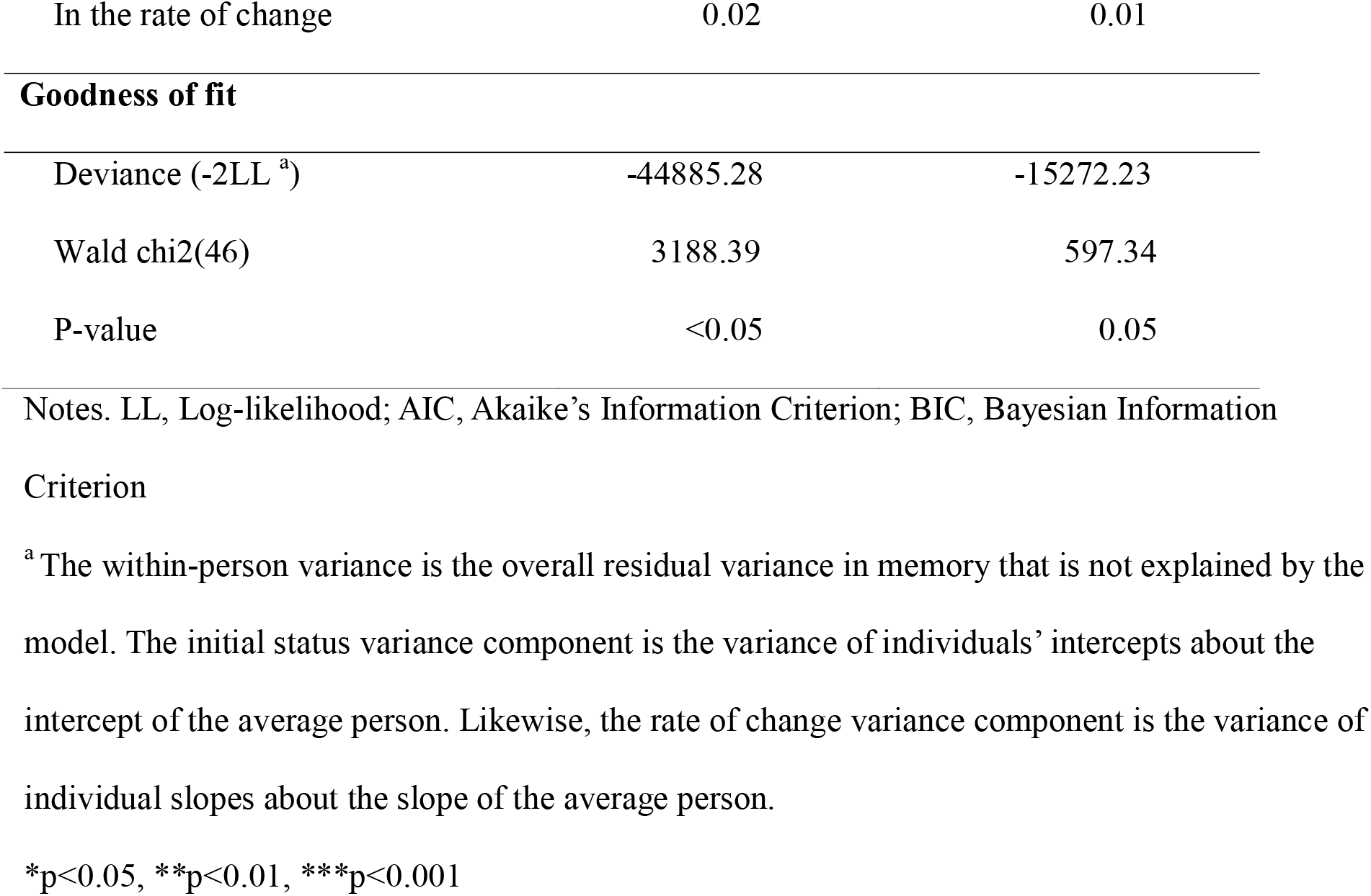
Linear mixed model of Vitamin D predicting the level of cognitive function and its change over time, 6 years of follow-up, N= 5365.

|  | Outcome: memory | Outcome: orientation |
| --- | --- | --- |
|  | Estimate (SE) | Estimate (SE) |
| **Initial Status** |  |  |
| Intercept | 7.45 (0.25) *** | 3.61 (0.04) *** |
| High Vitamin D level $\geq 31$ | Reference | Reference |
| Low Vitamin D level $< 31$ | -0.28 (0.11) * | -0.02 (0.02) |
| Age | -0.12 (0.01) *** | -0.01 (0.01) *** |
| Sex (female vs. male) | 1.10 (0.08) *** | 0.06 (0.01) *** |
| Unmarried vs married | 0.06 (0.09) | 0.03 (0.01) * |
| Education |  |  |
| Intermediate vs. no qualification | 0.95 (0.11) *** | 0.03 (0.02) * |
| Higher vs.no qualification | 1.68 (0.12) *** | 0.07 (0.02) *** |
| Wealth |  |  |
| 2<sup>nd</sup> quintile vs. lowest (1<sup>st</sup>) quintile | 0.39 (0.14) ** | 0.01 (0.02) |
| 3<sup>rd</sup> quintile vs 1<sup>st</sup> quintile | 0.57 (0.14) *** | 0.05 (0.02) * |
| 4<sup>th</sup> quintile vs 1<sup>st</sup> quintile | 0.90 (0.15) *** | 0.05 (0.02) * |
| 5<sup>th</sup> quintile vs. 1<sup>st</sup> quintile | 1.17 (0.15) *** | 0.07 (0.02) ** |
| Smoker vs. non-smoker | -0.18 (0.13) | -0.03 (0.02) |
| Physical exercise vs. no physical exercise | 0.46 (0.12) *** | 0.03 (0.02) |
| Waist |  |  |
| Medium vs Low | -0.01 (0.11) | 0.01 (0.02) |
| High vs. Low | -0.03 (0.10) | 0.03 (0.02) |
| Mobility impairment (yes vs. no) | -0.04 (0.02) * | -0.01 (0.01) ** |
| Coronary Heart Disease (CHD) (yes vs.no) | -0.23 (0.15) | -0.03 (0.02) |
| Stroke vs. No stroke | -0.37 (0.21) | 0.04 (0.03) |
| Diabetes vs. No diabetes | -0.18 (0.13) | -0.01 (0.02) |
| Hypertension vs. No hypertension | 0.10 (0.08) | 0.02 (0.01) |
| Depression vs. No depression symptoms | -0.5 (0.13) *** | -0.09 (0.02) *** |
| Season |  |  |
| Spring vs. Winter | -0.06 (0.16) | 0.01 (0.03) |
| Summer vs. Winter | -0.24 (0.11) * | -0.03 (0.02) |
| Autumn vs. Winter | -0.02 (0.10) | 0.01 (0.02) |
| **Rate of change** |  |  |
| Intercept | -0.26 (0.05) *** | -0.03 (0.01) ** |
| Vitamin D level <31 vs Vit. D level ≥31 | -0.01 (0.02) | 0.01 (0.01) |
| Age | -0.01 (0.01) *** | -0.01 (0.01) *** |
| Sex (female vs. male) | 0.01 (0.02) | -0.01 (0.01) |
| Unmarried vs married | -0.01 (0.02) | -0.01 (0.01) |
| Education |  |  |
| Intermediate vs. no qualification | 0.07 (0.02) ** | 0.01 (0.01) |
| Higher vs. no qualification | 0.07 (0.02) ** | 0.01 (0.01) |
| Wealth |  |  |
| 2<sup>nd</sup> quintile vs. lowest (1<sup>st</sup>) | -0.02 (0.03) | 0.01 (0.01) |
| quintile |  |  |
| 3<sup>rd</sup> quintile vs 1<sup>st</sup> quintile | -0.01 (0.03) | -0.01 (0.01) |
| 4<sup>th</sup> quintile vs 1<sup>st</sup> quintile | -0.02 (0.03) | 0.01 (0.01) |
| 5<sup>th</sup> quintile vs. 1<sup>st</sup> quintile | 0.03 (0.03) | 0.01 (0.01) |
| Smoker vs. non-smoker | -0.01 (0.03) | -0.01 (0.01) |
| Physical exercise vs. no physical exercise | 0.04 (0.02) | 0.02 (0.01) ** |
| Waist |  |  |
| Medium vs Low | 0.01 (0.02) | 0.01 (0.01) |
| High vs. Low | 0.02 (0.02) | 0.01 (0.01) |
| Mobility impairment (yes vs. no) | 0.01 (0.01) | 0.01 (0.01) |
| Coronary Heart Disease (CHD) (yes vs.no) | -0.04 (0.03) | 0.01 (0.01) |
| Stroke vs. No stroke | -0.01 (0.05) | -0.03 (0.01) * |
| Diabetes vs. No diabetes | -0.04 (0.03) | -0.01 (0.01) |
| Hypertension vs. No hypertension | 0.02 (0.02) | -0.01 (0.01) |
| Depressive symptoms vs. No depressive symptoms | -0.01 (0.03) | 0.01 (0.01) |
| Season |  |  |
| Spring vs. Winter | -0.01 (0.03) | -0.01 (0.01) |
| Summer vs. Winter | -0.04 (0.03) | -0.01 (0.01) |
| Autumn vs. Winter | -0.03 (0.02) | -0.01 (0.01) |
| Variance <sup>a</sup> |  |  |
| Within-person | 4.79 | 0.24 |
| In the initial status | 4.57 | 0.01 |
| In the rate of change | 0.02 | 0.01 |
| **Goodness of fit** |  |  |
| Deviance (-2LL <sup>a</sup>) | -44885.28 | -15272.23 |
| Wald chi2(46) | 3188.39 | 597.34 |
| P-value | <0.05 | 0.05 |

Notes. LL, Log-likelihood; AIC, Akaike's Information Criterion; BIC, Bayesian Information Criterion
<sup>a</sup>The within-person variance is the overall residual variance in memory that is not explained by the model. The initial status variance component is the variance of individuals' intercepts about the intercept of the average person. Likewise, the rate of change variance component is the variance of individual slopes about the slope of the average person.
\*p<0.05, \*\*p<0.01, \*\*\*p<0.001

### Blood levels of 25(OH)D and change in orientation scores over time

The associations between 25(OH)D blood levels and changes in orientation performance over time are presented in Table 2. The results of the fully adjusted model indicate that ELSA participants had a mean intercept of 3.61, SE=0.04, p≤0.001 in orientation. This score declined over time with an average rate of -0.03, SE=0.01, p≤0.01. Participants in the group with lower blood levels of 25(OH)D < 31nmol/l did not present lower baseline orientation scores in comparison with the reference group, i.e., blood levels of 25(OH)D ≥ 31nmol/l. Age (β= -0.01, SE=0.00, p≤0.001), mobility impairment (β= -0.01, SE=0.00, p≤0.01) and depressive symptoms (β= -0.09, SE=0.02, p≤0.001) suggested a negative association with orientation performance, while sex (being female, β= 0.06, SE=0.01, p≤0.001), being unmarried (β= 0.03, SE=0.01, p≤0.05), higher education (β= 0.07, SE=0.02, p≤0.001) and being wealthier (β= 0.07, SE=0.02, p≤0.01) were related to a better orientation.

There was no statistically significant association between blood levels of 25(OH)D and changes in orientation performance over time (β= 0.01, SE=0.01, p≥0.05). Among the covariates, age (β= -0.01, SE=0.01, p≤0.001) and having a stroke (β= -0.03, SE=0.01, p≤0.05) were related to a steeper decline in orientation performance in time, while being physically active (β= 0.02, SE=0.01, p≤0.01) was associated with a slower decline in orientation.

### Discussion

The present study examined the relationship between serum levels of 25(OH)D and changes over time in two domains of cognitive performance, i.e., memory (immediate and delayed) and orientation, in older English adults free of dementia at baseline from a long-running nationally representative sample of the English population. As hypothesized, our key findings showed a positive relationship between low circulating blood levels of 25(OH)D and low memory at baseline, in which a deficiency in serum levels of 25(OH)D was associated with poor baseline memory. On the other hand, there was no significant association between low blood levels of 25(OH)D and changes in memory performance after a 6-year follow-up. With regards to the performance in orientation, our results showed no significant association between low baseline blood levels of 25(OH)D and orientation neither at baseline nor change in orientation over time.

Our observations align with previous studies that reported positive associations between insufficient blood levels of 25(OH)D and lower scoring on one or more cognitive function tests (21). Previous studies suggested an association between insufficient levels of 25(OH)D and poor performance in tasks regarding episodic memory (22). Moreover, consistent with our observations, it has been found that lower blood levels of 25(OH)D were associated cross-sectionally with poorer scores on different memory tests but not significantly associated with the rate of change in memory performance in time (23). In accordance with our observations, previous studies could not find any specific association between serum levels of 25(OH)D and visual-spatial ability (24).

Cross-sectional and longitudinal studies conducted so far have reported conflicting results. While cross-sectional studies suggest that low levels of 25(OH)D might have a role in poorer cognitive performance, fewer longitudinal studies have been conducted to explore and confirm such findings (11). Moreover, there are only a few longitudinal studies in the current literature investigating the relationship between blood levels of 25(OH)D and changes over time in cognitive performance, particularly in a healthy population without any diagnosis of dementia. Longitudinal studies that explored the relationship between insufficient blood levels of 25(OH)D and more specific neuropsychological domains, such as different types of memory or visuospatial ability, have reported mixed results and need further investigations (25).

Most studies conducted so far have been predominantly cross-sectional, while the few longitudinal had used smaller population sizes. Our analysis is one of the first studies to examine the relationship between blood levels of 25(OH)D and cognitive performance in such a large population and over a long follow-up period, i.e., over 6 years. Moreover, it is one of the first longitudinal studies involving community-dwelling older English adults without dementia at baseline.

The inconsistency of the obtained results so far could be attributed to different factors. The heterogeneity of the studies makes it difficult to compare our findings. The variables included in the analysis of various studies, such as cognitive performance and blood levels of 25(OH)D, have often been assessed through heterogeneous techniques and diverse definitions. For example, different studies use different thresholds to determine what is an insufficient level of 25(OH)D and use various cognitive tests for measuring different domains of cognitive functioning. The current research, being among the first using the ELSA population sample, has used the same threshold of another previous study in ELSA to make our findings more comparable. Moreover, most authors consider blood levels of 25(OH)D under 30nmol/l to be deficient (18). Therefore, in the present analysis, the levels of blood concentration of 25(OH)D less than 31nmol/l were considered insufficient. The participants were divided into two groups: one with the participants who presented low blood levels of 25(OH)D < 31nmol/l and those with high blood levels of 25(OH)D ≥ 31nmol/l. Regarding the assessment of cognitive performance, only two domains were used. These were memory, which included immediate and delayed recall and orientation.

From all the covariates considered, being female, having a higher level of education, a higher socioeconomic status (being wealthier), and doing more physical activity were associated with better memory performance. Regarding the changes in time of memory performance, only education has been shown to be positively associated with slower memory performance decline over time. On the other hand, only age has been related to a faster memory decline. Examining the role of covariates on orientation, age, mobility impairment, and depression were associated with a poorer orientation at baseline, while being female, being unmarried, having a higher level of education, and having a higher socioeconomic status was associated with a better orientation performance at baseline. In reference to changes in time, age and having a stroke were related to a steeper decline in orientation performance in time, while physical activity was associated with a slower decline in time at orientation testing.

The positive association between blood levels of 25(OH)D and memory performance at baseline found in the present study agrees with the results from a few studies exploring the biological mechanisms involved. Previous studies have suggested a neuroprotective role of 25(OH)D by being involved in neurogenesis through nerve growth factor release, particularly in the hippocampus, which has a crucial role in memory (14). Further studies focusing on the long-term effects of insufficient 25(OH)D blood levels and changes in memory over time are needed, particularly focusing on the optimal timing and duration in which an increase in blood levels of 25(OH)D could play a protective role against memory decline. Moreover, the most recent developments in the field of neurobiology and neuroscience should be integrated into further research. For example, studies suggested that 25(OH)D has different variations in the receptor that might have other biological effects on how an insufficient blood level of 25(OH)D could lead to various chronic diseases (26).

### Strengths and limitations

The current analysis has several strengths and limitations. Among the strengths of the study are its large sample size, representative of the English population aged 50 and older, and a relatively long follow-up of six years. Serum samples of 25(OH)D have been collected objectively in laboratories, following standardized protocols, detecting both 25(OH)D_2_ and 25(OH)D_3_, allowing an analysis of the total circulating of 25(OH)D. Another strong point relates to methodology and the inclusion of a wide range of covariates. Linear Mixed Model is an optimal choice to analyze changes in continuous dependent variables (memory and orientation scores) over time and to evaluate the association with a diverse range of covariates. A particular advantage is that mixed linear models prevent false-positive associations due to these types of systems and provide an increased power specific to the sample structure. Even though the mixed model analyses use maximum likelihood estimation to account for attrition over the 6-year follow-up period, the population sample may be initially selected on critical variables. Those with more education, better financial circumstances, and better cognitive performance were more likely to accept the initial invitation to participate in the study. Lastly, to avoid practice effects, the memory assessments employed by ELSA used alternative lists of words at each wave and have undergone monthly quality control to check for inter and intra-rater reliability. Furthermore, the 2-year gap between memory assessments in ELSA’s design indicates a reduced possibility of retest effects, which is common in longitudinal studies of cognitive aging.

However, there are a number of limitations that should be acknowledged. One of the limitations of this observational study is that we cannot determine causality in the relationship between blood levels of 25(OH)D and cognitive performance. The possibility of reverse causality in interpreting the results must be considered. For example, it might be possible that the participants with better cognitive function in memory and orientation are more likely to do more physical exercise outdoors, have more sun exposure, eat healthier (because they can afford to have a higher socioeconomic status and the knowledge to do so) and therefore will consequently present higher circulating 25(OH)D. To address this limitation, further intervention studies need to be conducted. In the current literature, only a few intervention studies have been conducted so far, with inconclusive results (11).

Moreover, ELSA does not have any additional information on the differences between 25(OH)D_2_ and 25(OH)D_3._ When exposed to sunlight, the blood levels of 25(OH)D_3_ increase, whilst the blood levels of 25(OH)D_2_ are influenced mainly by diet (27). Almost half of the participants of the current analysis, 43.45%, have had their sample collected in autumn, which might have had some implication regarding the actual 25(OH)D levels. For example, during autumn, participants might have less exposure to the sun, presenting consequently lower blood levels of 25(OH)D_3_. Knowing the difference between 25(OH)D_2_ and 25(OH)D_3_, investigating what factors influence their levels and how their blood levels impact cognitive performance would help future research in setting up more accurate interventions.

Another limitation of the study is that 97% of the ELSA participants identified themselves as “white English”, highlighting a lack of ethnic diversity in this study. This limits the generalisability of the results as it is possible that ethnicity may play a role as a mediator or act as a covariate in the relationship between serum levels of 25(OH)D and cognitive performance, and therefore these results should be interpreted with caution.

In interpreting the results, two possible biases should be considered: attrition and selection bias. First, due to the longitudinal design of the study, attrition could lead to an attrition bias. Second, the possibility of selection bias must be acknowledged, as people who participated in the study might differ from the general population.

Lastly, another limitation of this study relates to the assessment of cognitive performance. Cognition is a broad and general concept that comprises lots of domains to be assessed. Measuring only orientation and memory might be too restrictive to generalize the results to overall cognitive performance.

## Conclusions

In summary, this epidemiological study based on a nationally representative sample of older English adults showed a significant association between low serum 25(OH)D concentrations with poorer memory performance at baseline but did not find evidence for an association with changes in cognitive performance (either memory or orientation) over time. The findings suggest that 25(OH)D blood levels might play a role in memory performance, contributing to the current literature that has found a similar effect.

However, further research is necessary to understand if our findings have any clinical implications and whether the results could be used to implement interventions to improve public health. Randomized control trials are needed to establish a causal relationship between 25(OH)D and cognitive decline. At the same time, more longitudinal studies focusing on 25(OH) blood levels and expanding the assessment of cognitive performance will provide further clarification in understanding the role of 25(OH)D on cognitive decline over time.

## Data Availability

The English Longitudinal Study of Ageing (ELSA) was developed by a team of researchers based at University College London, the Institute for Fiscal Studies, and the National Centre for Social Research. The data are linked to the UK Data Archive and freely available through the UK data services and can be accessed here: https://discover.ukdataservice.ac.uk.

## Declarations

### Ethics approval and consent to participate

Ethical approval for each of the ELSA waves was granted by the National Research Ethics Service (London Multicentre Research Ethics Committee). All participants gave informed consent. All methods and the study are GDPR compliant.

### Author contribution

SG and DC were responsible for data analysis and drafting the report. DC was responsible for the study conception and design, preparation of data, data analysis, and the critical revision of the report. SG was responsible for study conception and design, preparation of data, data analysis, and drafting the first draft of the manuscript. All authors were responsible for the interpretation of the results of critical revision, and all approved the final version of the report.

## Acknowledgements

The authors express gratitude to the ELSA research participants and the professionals involved in conducting the study, making it an invaluable source of information for researchers around the world.

